# Genetically Determined Blood Pressure, Antihypertensive Drugs, and Genes with Clinical Outcome after Ischemic Stroke: Evidence from Mendelian Randomization

**DOI:** 10.1101/2023.04.09.23288342

**Authors:** Hanchen Liu, Xiaoxi Zhang, Yu Zhou, Thanh N. Nguyen, Lei Zhang, Pengfei Xing, Zifu Li, Hongjian Shen, Yongxin Zhang, Weilong Hua, Hongye Xu, Xuan Zhu, Chen Lei, Qiao Zuo, Rui Zhao, Qiang Li, Dongwei Dai, Yongwei Zhang, Yi Xu, Qinghai Huang, Jianmin Liu, Pengfei Yang

## Abstract

**Background:** Observational studies suggest a robust association between blood pressure (BP) and functional outcomes in ischemic stroke patients. We sought to identify novel associations of the genetic basis of 3-month functional outcome after ischemic stroke based on a Mendelian randomization (MR) framework.

**Methods:** We selected genetic variants associated with systolic and diastolic BP and BP-lowering variants in genes encoding antihypertensive drugs from genome-wide association studies (GWAS) on 757,601 individuals. The primary outcome was 3- month favorable functional outcome defined as modified Rankin Scale (mRS) of 0-2. The secondary outcome was excellent 90-day outcome defined as mRS 0-1. The Cochran’s Q statistic in Inverse variance weighted (IVW) model, the weighted median, MR-Egger regression, leave-one-SNP-out analysis, MR-Pleiotropy Residual Sum and Outlier methods were adopted as sensitivity analyses. To validate our primary results, we performed independent repeat analyses and Bi-directional MR analyses.

**Results:** Genetic predisposition to higher systolic and diastolic BP was associated with a lower probability of 3-month excellent functional outcome after ischemic stroke in univariable IVW MR analysis (OR=1.29, 95%CI 1.05-1.59, *p*=0.014; OR=1.27, 95%CI 1.07-1.51, *p*=0.006, respectively). Pulse pressure was associated with both excellent and favorable functional outcome (OR=1.05, 95%CI 1.02-1.08, *p*=0.002; OR=1.04, 95%CI 1.01-1.07, *p*=0.009, respectively). Angiotensin-converting enzyme inhibitor (ACEI) and calcium channel blocker (CCB), were significantly associated with improved favorable functional outcome (OR=0.76, 95%CI 0.62-0.94, *p*=0.009; OR=0.89, 95%CI 0.83-0.97, *p*=0.005). Proxies for β-blockers, angiotensin receptor blocker (ARB) and thiazides failed to show associations with functional outcome (*p*>0.05).

**Conclusion:** We provide evidence for a potential association of genetic predisposition to higher BP with higher risk of 3-month functional dependence after ischemic stroke. Our findings support ACEI and CCB as promising antihypertensive drugs for improving functional outcome in ischemic stroke.

## Introduction

High blood pressure (BP) is commonly seen in ischemic stroke patients^(1-3)^ and a genetic predisposition to higher BP has been associated with an increased risk of any stroke, ischemic stroke, and its main subtypes.^(4)^ Randomized controlled trials (RCTs) found that BP lowering treatment with anti-hypertensive drugs including ACEI, ARB and CCB is beneficial in lowering stroke risk.^(5, 6)^ Pre-stroke use of antihypertensive drugs is also related with better clinical outcome.^(7)^ However, it remains unknown whether genetic predisposition leads to less severe stroke or better clinical outcome after ischemic stroke onset.

Mendelian randomization (MR) uses single nucleotide polymorphisms (SNPs) as proxies for traits of risk factors and is less prone to confounding bias and reverse causation than observational studies.^(8)^ Of note, MR is commonly considered valuable in exploring causality and in predicting the effect of interventions, which may explain possible intervention targets in clinical practice including antihypertensive treatment.^(9-11)^ Genome-wide association studies (GWAS), with large samples and genetic data, further permit prediction of clinical outcomes of ischemic stroke, especially in the absence of data from RCTs. Simultaneously, the effects of drug action can be anticipated by the genetic effects in the genes of their protein targets, as has previously been applied to antihypertensive drugs. ^(4)^

Herein, we employed MR to examine the effect of genetically determined BP and its proxies for antihypertensive drugs on clinical outcome in ischemic stroke, with data from the UK Biobank and International Consortium of Blood Pressure of European ancestry.

## Methods

### Data Availability

This study was conducted in accordance with the guidelines for Strengthening the Reporting of Observational Studies in Epidemiology-Mendelian randomization (STROBE-MR).^(12)^ We used summary data from published studies for our analyses, publicly available via the original studies. All studies obtained relevant ethical approval and participant consent.

### Exposure Data

Genetic instruments for systolic BP (SBP), diastolic BP (DBP) and pulse pressure (PP) were obtained from the summary statistics of the GWAS meta-analysis consisting of 757,601 individuals (458,577 from the UK Biobank and 299,024 from the International Consortium of Blood Pressure) of European ancestry.^(13)^ In the pooled sample, mean SBP and DBP were 138.4 (SD 21.5) and 82.8 (SD 11.4) mm Hg, and the mean age of participants ranged from 56.8 to 62.1 years old. As genetic instruments for SBP and DBP, we selected single nucleotide polymorphisms (SNPs) associated with SBP, DBP, and PP at genome-wide significance level (P < 5 × 1 0^−8^) and clumped for linkage disequilibrium (LD) to r^2^ < 0.001 based on the European 1,000 Genomes panel.

Five commonly used antihypertensive drugs were selected, including ACEI, ARB, BB, CCB and thiazide diuretics, according to guidelines.^(14)^ Genetic instruments for the effects of antihypertensive drugs were obtained as the variants in the drug targeted genes associated with BP at a genome-wide significant level ^(4, 15, 16)^. Genes encoding the pharmacologic targets related to the effect of common antihypertensive drugs on

BP were identified in DrugBank.^(17)^ After, the genomic regions and their regulatory regions (promoters and enhancers) corresponding to these genes were screened in Genecards.^(18)^ From all the identified variants in each gene, only variants that were significantly associated with SBP (P < 5 × 1 0^−8^) and clumped to a LD threshold of r^2^ < 0.4 using the 1000G European reference panel were considered as candidate proxies for each medication class. This relatively lenient LD threshold allows for an increase in proportion of variance explained and thus in statistical power ^(19, 20)^. For additional analysis, we also employed more stringent LD threshold (r^2^ < 0.1). There is the only one genetic variant for ACEI. To improve statistical capabilities, another method was used to explore construction of genetic instruments to proxy ACEI: by obtaining genome-wide significant variants in week linkage disequilibrium (r^2^ < 0.1) in or within ±100 kb from angiotensin-converting enzyme (ACE) that were associated with serum ACE concentrations in a GWAS of 4, 174 participants in the Outcome Reduction with Initial Glargine Intervention study (resulting in 14 SNPs).^(21)^ These proxies for ACEI have been validated in previous study. ^16^

### Outcome Data

The primary outcome was favorable functional outcome, defined as mRS (modified Rankin scale) 0 to 2. The secondary outcome was excellent functional outcome defined as mRS 0-1. The mRS evaluates global disability with scores ranging from 0 (no symptoms) to 6 (death). A score of 2–5 indicates some degree of disability.(22) We obtained the summary-level data for functional outcome after ischemic stroke from the Genetics of Ischemic Stroke Functional Outcome (GISCOME) network meta-analysis, which contains 12 studies with 6,021 stroke patients from the US, Europe, and Australia.^(23)^ In the primary analysis, we used the summary data which was adjusted for age, sex, principal components, and baseline NIHSS score. In the repeated analysis, we used the data without adjustment for baseline NIHSS to verify our primary results.^(23)^ All individuals were of European ancestry.

### Statistical Analysis

We conducted primary MR analysis using a random-effects IVW meta-analysis to explore the associations for BP and antihypertensive medications with functional outcome after ischemic stroke. IVW MR methods assumes that all variants are valid instruments^(24)^ and comprises a meta-analysis of SNP-specific Wald estimates 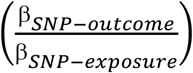 using an inverse of the corresponding variance.^(25)^ We excluded the SNPs that had F statistics lower than 10 according to standard practice.^(26)^ All MR associations between SBP, DBP, PP, and primary outcomes were scaled to 10 mm Hg increment in SBP, 5 mm Hg in DBP, and 1 mm Hg in PP.

We performed several sensitivity analyses. First, we used Cochran’s Q statistic in the IVW model to assess the heterogeneity between variant-specific estimates. If possible, we used the results of the weighted median method. The weighted median approaches give more weight to more precise instrumental variables and the estimate is consistent even when up to 50% of the information comes from invalid or weak instruments.^(27)^ Afterwards, we used MR-Egger and MR-Pleiotropy Residual Sum and Outlier methods to explore horizontal pleiotropy.^(28)^ Finally, we conducted leave-one-SNP-out analysis, in which SNPs were systematically removed, to assess if results were driven by single SNP. The association is considered significant after correction for multiple testing for three BP indexes [P < 0.016 (0.05/3)] and five antihypertensive medications’ classes [P < 0.01 (0.05/5)]. We calculated all effect estimates as adjusted odds ratios (OR) with 95% CIs. The study design is shown in **Figure 1**.

**Figure 1.**
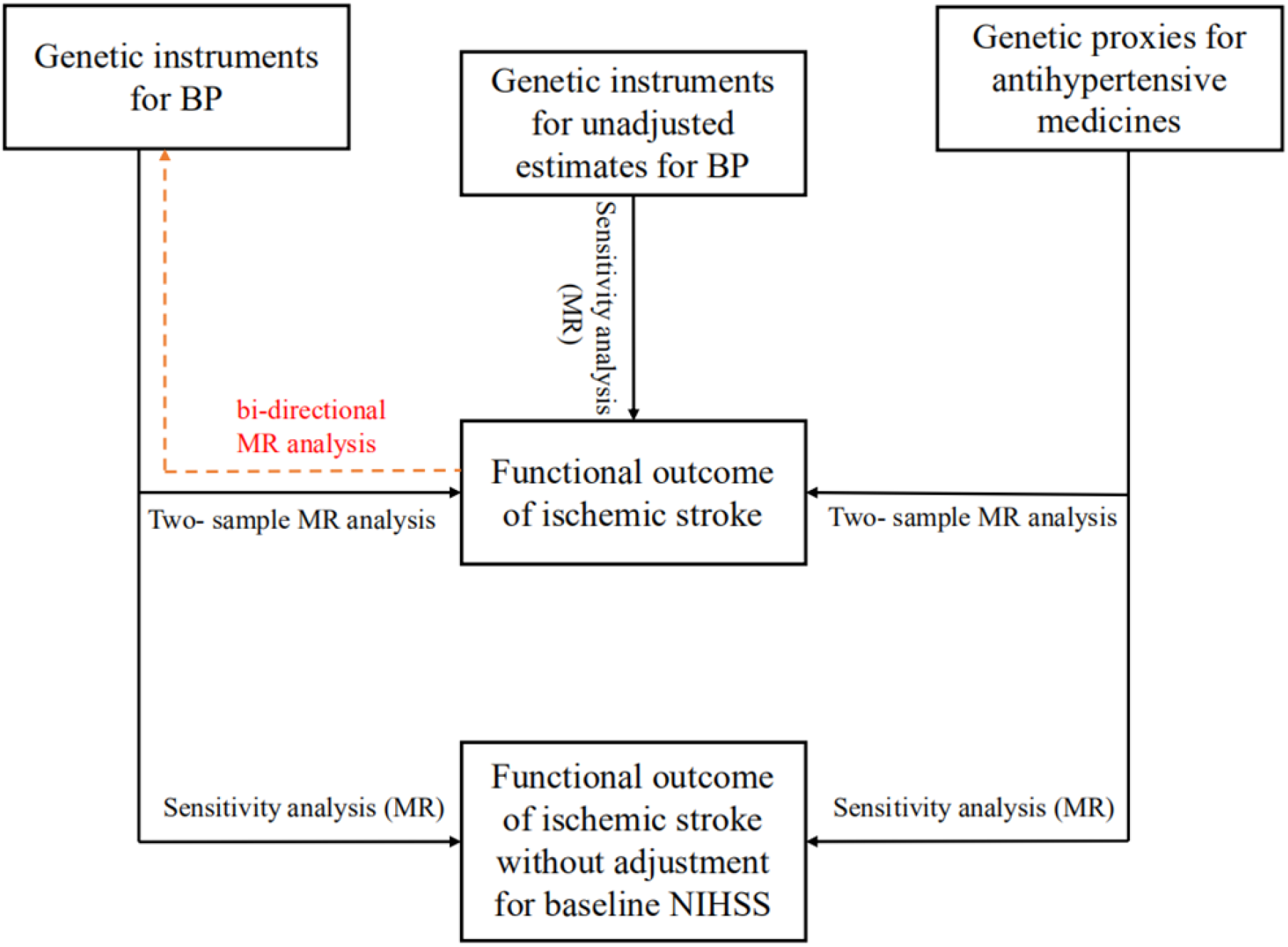
Flowchart Depicting Study Design.

To validate the primary results, we repeated the analyses in the outcome database without adjustment for baseline NIHSS and estimated the effects of antihypertensive drugs on functional outcome after ischemic stroke by selecting SNPs that were at more stringent LD threshold (r2 < 0.1). To eliminate potential bias due to medication noncompliance or collider effects, we performed sensitivity analyses using unadjusted estimates for BP from a UKB GWAS (317,756 individuals).^(29)^ Moreover, we included a bi-directional MR analysis to explore whether functional outcome after ischemic stroke affects to BP. All analyses were performed via Two Sample MR (version 0.5.6), Mendelian randomization (version 0.5.1), and MRPRESSO (version 1.0) packages in R version 4.2.1.^(30)^

## Results

### Genetically determined BP and functional outcome after ischemic stroke

There were 368, 374 and 328 independent genetic variants verified to be associated with SBP, DBP and PP, respectively (**Table S1-S4**). **Figure 2** illustrates the result of the primary univariable MR analysis. A 10 mmHg increase of genetically determined SBP was associated with decreased probability of excellent functional outcome after ischemic stroke (OR 1.29, 95% CI: 1.05-1.59, P = 0.014). A 5 mmHg increase of DBP was associated with a decreased probability of excellent functional outcome (OR 1.27, 95% CI: 1.07-1.51, P = 0.006). Interestingly, each 1 mmHg increase of PP was associated with a decreased probability of both excellent and favorable clinical outcome (OR 1.05, 95% CI: 1.02-1.08, P = 0.002; OR 1.04, 95% CI: 1.01-1.07, P = 0.009). The lower likelihood of a favorable functional outcome with an increase of SBP and DBP, however, was insignificant (OR 0.91, 95% CI: 0.74-1.12, P = 0.358; OR 1.04, 95% CI: 0.89-1.23, P = 0.607, respectively).

**Figure 2.**
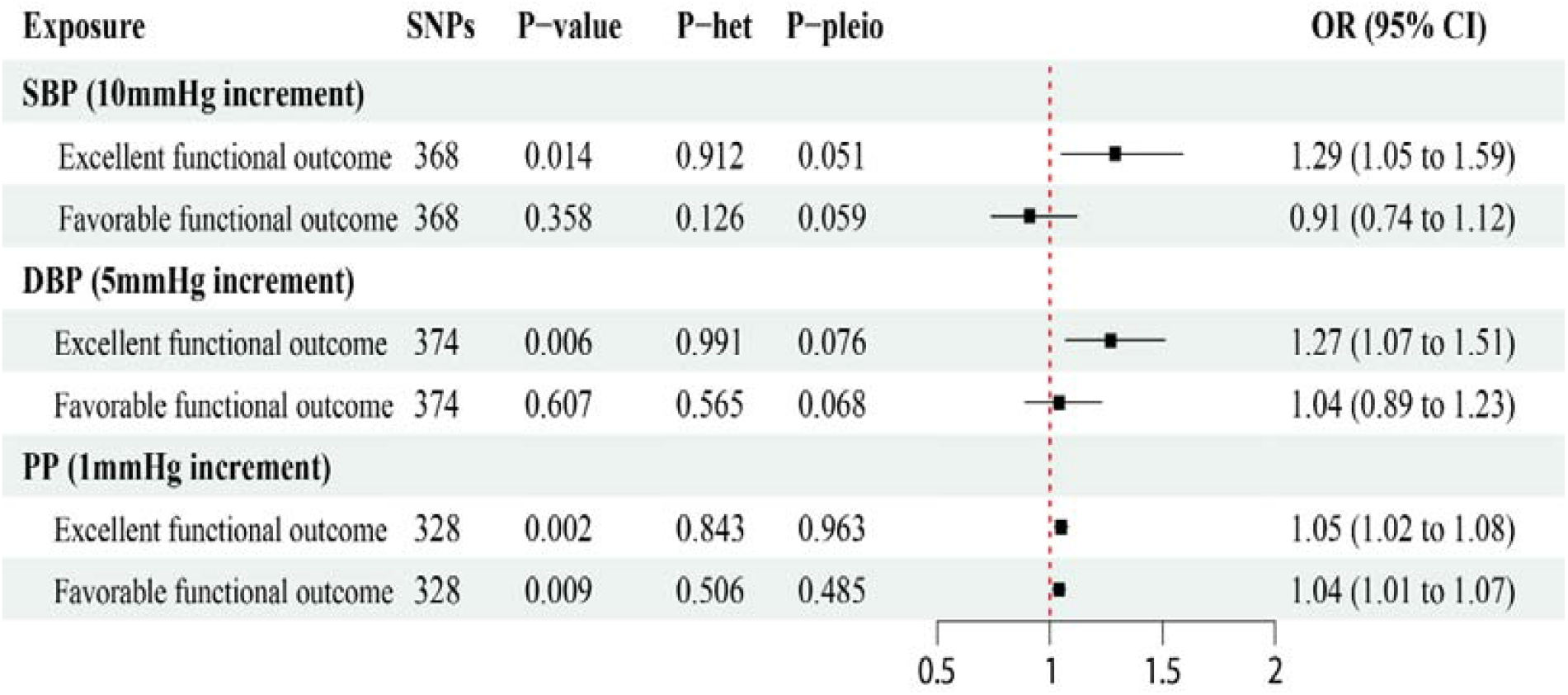
Genetically predicted blood pressure with functional outcome after ischemic stroke. Genome-wide significantly associated (P < 5 × 10^−8^) independent (LD R^2^ = 0.001, clumping distance = 10,000 kb) SNPs were used as instruments. MR, Mendelian randomization; SNP, single nucleotide polymorphism; OR, odds ratio; CI, confidence interval; SBP, systolic blood pressure; DBP, diastolic blood pressure; PP, pulse pressure; IA, Intracranial aneurysm; SAH, Subarachnoid Hemorrhage; P-het, P value in the Q statistic for heterogeneity; P-pleio, P value in the Egger intercept. P value in the Q statistic for heterogeneity; P-pleio, P value in the Egger intercept.

No evidence for heterogeneity was verified in the Cochran Q test and no directional pleiotropic effects were observed in MR-Pleiotropy Residual Sum, Outlier global test and MR-Egger intercepts. Results from the weighted median method also supported a positive association, which was not significant (**Table S5**). Leave-one-SNP-out analysis showed that results were robust with all SNPs and were not driven by any single SNP (**Figure S1**).

### Genetic proxies for antihypertensive drugs and functional outcome after ischemic stroke

BP-lowering variants in genes encoding drug targets were tested as proxies for the effects of antihypertensive drugs and their effects on clinical outcomes. A total of 98 variants for antihypertensive drugs, including 14 for ACEI, 11 for ARB, 30 for BB, 38 for CCB, and 5 for thiazides were identified (**Table S4**). As shown in **Figure 3**, a genetically determined effect of ACEI was positively associated with a protective effect on favorable functional outcome after ischemic stroke (OR 0.76; 95% CI: 0.62-0.94, P = 0.009). A genetically determined effect of CCB also had a protective role (OR = 0.89; 95%CI: 0.83-0.97, P = 0.005). However, there was no causal effect of ARB, BB, and thiazide on functional outcome after ischemic stroke.

**Figure 3.**
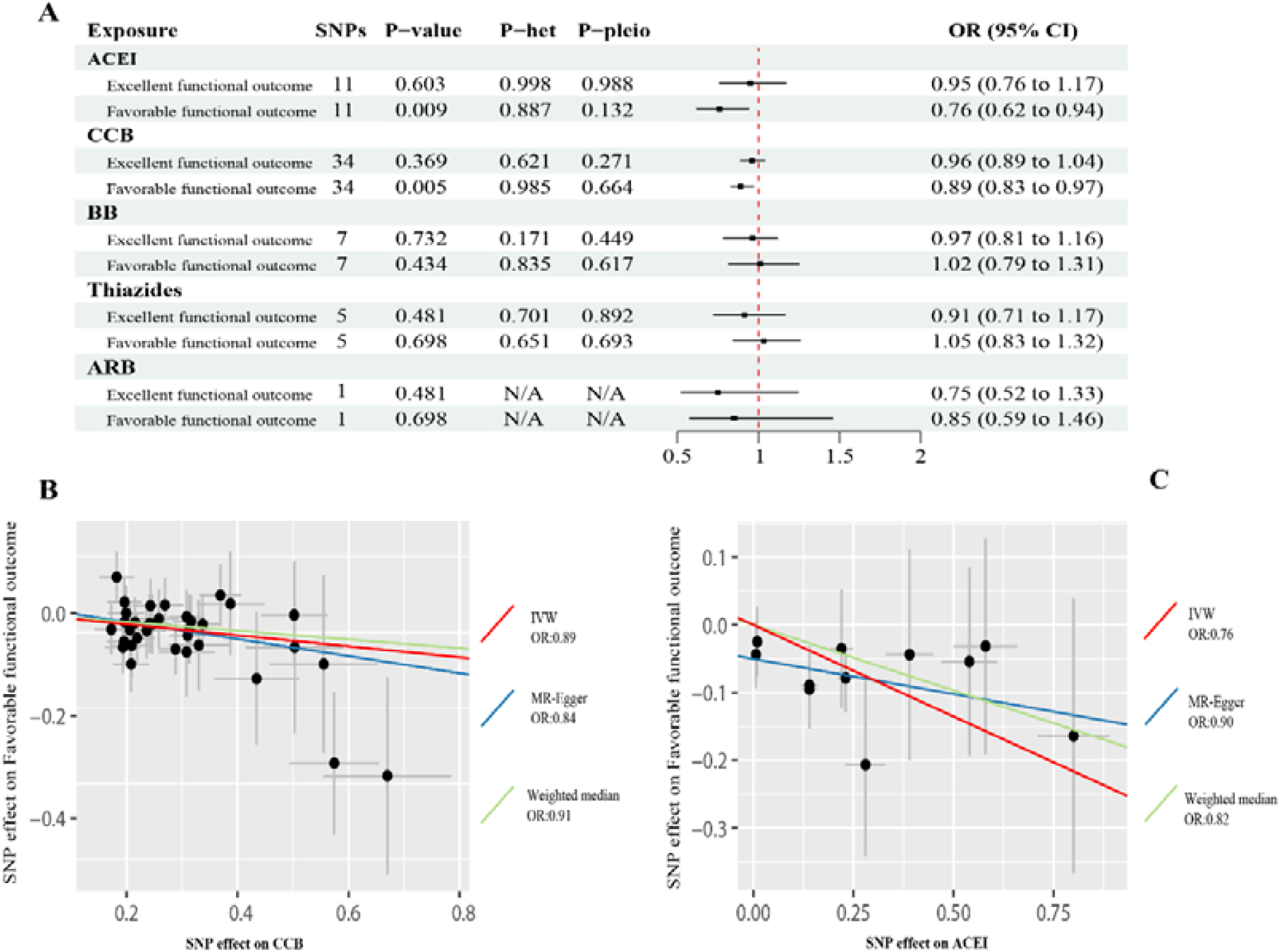
Genetically predicted antihypertensive drugs with functional outcome after ischemic stroke. A, Inverse-variance weighted estimates for the association between antihypertensive drugs on the functional outcome after ischemic stroke. B and C, Scatter plots of individual single-nucleotide polymorphism (SNP) effects and estimates from different Mendelian randomization (MR) methods for the effect of (B) calcium channel blocker (CCB) on Favorable functional outcome and (C) angiotensin-converting enzyme inhibitor (ACEI) on Favorable functional outcome. MR, Mendelian randomization; SNP, single nucleotide polymorphism; OR, odds ratio; CI, confidence interval; ACEI: angiotensin-converting enzyme inhibitor; ARB: angiotensin receptor blocker; CCB: calcium channel blockers; BB: beta-blockers; ACEI: angiotensin-converting enzyme inhibitors; ARB: angiotensin receptor blockers; P-het, P value in the Q statistic for heterogeneity; P-pleio, P value in the Egger intercept.

The Cochran Q statistic of the IVW method indicated no horizontal pleiotropy or heterogeneity across instrument SNP effects (**Figure 3**). Additional analysis restricted to the set of SNPs with the LD threshold (R2 < 0.1) showed consistent association estimates with primary results. A MR-Egger analysis did not show evidence of directional pleiotropy (P > 0.05). The results of the weighted median method also supported a protective effect of both drugs (**Table S6**). There was no distortion in the leave-one-out plot, suggesting that no single SNP was driving the observed effect in any analysis (**Figure S1**).

### Supplementary analyses

To validate our results, we performed the repeated analyses in the outcome database without adjustment for baseline NIHSS. The MR analysis showed results consistent with the primary analyses that a genetic predisposition to hypertension was associated with worse functional outcome after ischemic stroke (1 mmHg increment in SBP, mRS 0-1 vs 2-6, OR 1.02; 95%CI: 1.01-1.05, P = 0.009), (1 mmHg increment in DBP, mRS 0-1 vs 2-6, OR 1.05; 95%CI: 1.01-1.08, P = 0.005), (1 mmHg increment in PP, mRS 0-1 vs 2-6, OR 1.04; 95%CI: 1.02-1.07, P = 0.007), (ACEI, mRS 0-2 vs 3-6, OR 0.81; 95%CI: 0.68-0.97, P = 0.02), (CCB, mRS 0-2 vs 3-6, OR 0.91; 95%CI: 0.84-0.96, P = 0.002).

Next, sensitivity analyses using unadjusted estimates for BP were conducted to further verify the primary results. MR analysis results showed a 91% and 66% increased risk of worse functional outcome after ischemic stroke per 1-SD increase in genetically determined SBP (1-SD increment in SBP, mRS 0-1 vs 2-6, OR = 1.90; 95%CI: 1.11-3.26, P = 0.01) and DBP (1-SD increment in DBP, mRS 0-1 vs 2-6, OR = 1.66; 95%CI: 1.02-2.71, P = 0.042), respectively.

Finally, the result of the bi-directional MR analysis showed that there was no evidence that functional outcome after ischemic stroke is related to BP. Sensitivity analysis of all supplementary studies revealed no heterogeneity or pleiotropy in the replication analysis. (**Table S5-S6**)

## Discussion

We investigated the genetic relationship between BP variables, antihypertensive drugs, SNP with 3-month clinical outcome of ischemic strokes by leveraging large-scale genetic data including the GWAS and UKB data. We found a genetic predisposition to higher systolic and diastolic BP was associated with worse 3-month functional outcome after ischemic stroke. Prior use of ACEI and CCB, but not ARB, β-blockers or thiazides, were significantly associated with improved functional outcome after ischemic stroke after adjusting for confounders.

Predisposition to higher BP has been associated with higher risk of ischemic and hemorrhagic stroke at a genetic level.^(4)^ *Georgakis et al*^*(4)*^ performed MR investigating the genetic effects of BP and BP-lowering through different antihypertensive drug classes on stroke risk, and found that a genetic predisposition to higher BP was associated with risk of ischemic and hemorrhagic stroke. They also found that CCBs, but not β-blockers, were associated with lower ischemic stroke risk. Proxies for CCBs exhibited a strong association with small vessel disease and white matter hyperintensity phenotype, underscoring CCB as a promising strategy for the prevention of cerebral SVD. Our findings are partially consistent with the above result. We found a beneficial effect of ACEIs as well as a neutral effect of ARB and thiazides on functional outcomes after ischemic stroke. It is unclear whether ACEI and CCB use before stroke provides a protective effect resulting in less severe stroke. The mechanisms of a beneficial effect of ACEIs on reduced risk of ischemic stroke and improved functional outcomes are complex. The possible interpretations include a BP-lowering effect, anti-inflammatory effect independent of a BP-lowering effect, protection and reconstruction of endothelial cells.^(31-33)^ *Di Napoli et al*^*(34)*^ showed that ACEI administration at the time of ischemic stroke was associated with lower inflammatory response (manifest by reduced C-reactive protein) and better functional outcome, independent of its effects on BP.^(35)^ This phenomenon was also observed in several clinical trials. *Chitravas et al*^*(36)*^ assessed the effect of previous use of ACEI in 761 ischemic stroke patients and found a reduced risk of severe neurologic deficit (OR=0.56; 95% CI 0.35-0.91) and death within 28 days (OR=0.46; 95% CI 0.24-0.87). *Yahaya et al*^(37)^ performed a retrospective cohort study of 327 patients with acute ischemic stroke and 119 (36.4%) had documented previous ACEI administration. They found a lower in-hospital mortality risk among patients who were on ACEI before the index event (*p*=0.002). *Thomas et al*^*(38)*^ performed a meta-analysis of 11 studies and found that antihypertensive drugs were related with a lower risk of recurrent stroke in patients with a history of stroke or transient ischemic attack, and the result was primarily derived from trials studying an ACEI or a diuretic.

ARB was also shown to be beneficial in reducing the incidence of ischemic stroke and improving clinical outcome. A post-hoc analysis of the BP-TARGET trial found that patients treated with ARB before stroke exhibited less severe stroke and developed less intracranial hemorrhage at 24 hours.^(7)^ The potential mechanism could be explained by RAS blockage, which leads to decreased ICH.^(39)^ Patients taking a RAS inhibitor also tended to have more neurological improvement compared with the non-RAS inhibitor group, which could be partially explained by a higher rate of ICH in the group of patients who had no RAS inhibitor.^(7)^ Given the short half-life of antihypertensive drugs, there is the possibility that the biological effects of these baseline antihypertensive drugs only last a few hours during the acute phase, and thereafter, is insufficient to impact 3-month outcomes. In this MR analysis, previous use of ACEI showed a robust effect on improving 3-month clinical outcomes. We believe this signal suggests that ACEI in the acute phase of ischemic stroke could be beneficial in improving 3-month clinical outcomes, and would warrant further investigations. The opposite direction of the treatment effect of ACEI and ARB should be carefully interpreted. A nationwide study compared the risk of major adverse cardiac events between patients who initiated ACEI (n=15,959) or ARB (n=23,929) with type 2 diabetes.^(40)^ No difference was observed between the two groups. In contrast, another study showed that ARB prevented atrial fibrillation better than ACEI in patients with a history of prior stroke or TIA.^(41)^ In addition, the number of identified ARB proxies was 7, which may lead to an underestimated result of ARB on clinical outcome.

Key strengths of our study were the efforts to identify the causal relationship between blood bressure, antihypertensive drugs, and genes with clinical outcome on genetic level, with no bias or influence from the incidence of stroke. There are limitations of our study. First, MR examines the lifetime effect of genetically determined BP, antihypertensive drugs and BP related SNPs, which is different from the effect of blood pressure lowering treatment. The clinical effect of different antihypertensive medications needs validation in randomized clinical trials. Second, based on the limited baseline characteristics, we identified only the effect of BP on 3-month clinical outcome, which did not account for baseline stroke subtype and pre-stroke mRS scores. Further pre-clinical and clinical studies encompassing carefully selected participants are warranted to offer deeper insights into differential effects between classes of antihypertensive drugs, including ACEIs, ARBs, CCBs on ischemic stroke and stroke subtypes. Third, the genetic data were restricted to participants of European ancestry, which lead to low generalizability of our findings to other populations^(42)^. Large-scale GWAS and SNP data from a more diverse population with higher representation of non-European ethnicities could help to understand potential ethnic differences. Furthermore, the number of antihypertensive proxies was relatively small. Forth, it is pertinent that the relationship between blood pressure and clinical outcomes might be non-linear, for instance, a J- or U-shaped relationship^(43)^. Finally, our study evaluated stroke outcomes and did not account for stroke incidence or stroke recurrence. The interpretation of our results was restricted to overall ischemic stroke, without examining subgroup etiology.

We provide evidence for a potential association of a genetic predisposition to higher systolic and diastolic BP with worse 3-month functional outcome in patients with ischemic stroke. Our findings support ACEIs and CCBs, but not ARBs, β-blockers nor thiazides, to provide BP-lowering effect and improve 3-month functional outcome in ischemic stroke patients.

## Data Availability

All data mentioned in the manuscript are true and available

## Abbreviations

ACEI: angiotensin-converting enzyme inhibitor
ARB: angiotensin receptor blocker
AIS: acute ischemic stroke
OR: odds ratio
BP: blood pressure
CCB: calcium channel blocker
CI: confidence interval
DBP: diastolic blood pressure
GWAS: genome-wide association studies
ICH: intra-cranial hemorrhage
LD: linkage disequilibrium
IVW: Inverse variance weighted
MR: Mendelian randomization
mRS: modified Rankin Scale
PP: pulse pressure
RAS: renin-angiotensin system
RCT: randomized controlled trials
SD: standard deviation
SNP: single nucleotide polymorphisms
SBP: systolic blood pressure
SVD: small vessel disease

## Acknowledgment

Outcome data were accessed through the GISCOME network and ISGC Cerebrovascular Disease Knowledge Portal.

## Disclosure

The authors report no disclosures relevant to the manuscript. Go to Neurology.org/N for full disclosures.

### Supplemental Material

Tables S1-S7

Figure S1

STROBE-MR Checklist

